# Mind the Baseline: The Hidden Impact of Reference Model Selection on Forecast Assessment

**DOI:** 10.1101/2025.08.01.25332807

**Authors:** Manuel Stapper, Sebastian Funk

**Affiliations:** Department of Infectious Disease Epidemiology and Dynamics, London School of Hygiene & Tropical Medicine, London, UK

**Keywords:** Baseline models, COVID-19, epidemic forecasting, forecast evaluation, influenza, model comparison

## Abstract

Baseline models are essential reference points for evaluating forecasting methods, yet their selection often receives insufficient attention. We present a systematic framework for baseline model selection in epidemiological forecasting, establishing criteria for suitable baselines and demonstrating the consequences of different choices. Analysing data from COVID-19 and influenza forecast hubs, we evaluated ten baseline model frameworks. Our results reveal that baseline selection profoundly impacts forecast evaluation: for influenza, the proportion of models outperforming the baseline ranged from 11% to 100% depending on the baseline chosen. No single baseline satisfied all evaluation criteria. The choice of baseline also affected model rankings, with some baselines producing substantially different orderings of forecast model performance. We found that well-calibrated baselines do not necessarily align with good forecast performance, highlighting a fundamental tension in baseline selection. These findings highlight the need for careful baseline selection in forecast evaluation, particularly in collaborative efforts where fair comparison across multiple models is essential. We provide practical recommendations for baseline selection and suggest strategies for improving evaluation fairness when ideal baselines cannot be identified.

## 1 Introduction

Evaluating the performance of forecasting models is a cornerstone of predictive modeling in epidemiology and beyond. When a new forecasting method or model is proposed, researchers typically compare its performance with existing approaches to demonstrate its value. Central to this evaluation process is the choice of a baseline model: a simple reference model against which more sophisticated methods are compared. Despite its fundamental role in forecast evaluation, the selection of baseline models (sometimes referred to as null models) often receives insufficient attention, potentially leading to misleading conclusions about model performance.

A baseline model serves as a reference point for evaluating a single or multiple forecasting methods. It is typically a simple model that provides a standard of comparison, allowing researchers to quantify the added value of more complex approaches. For instance, when evaluating regression models in-sample, the overall mean serves as an implicit baseline through the *R*^2^, quantifying how much variance a model explains beyond simply predicting the average. In time series forecasting, common baselines include the “no-change” forecast (using the most recent observation) or seasonal averages from historical data. The choice of baseline can profoundly influence conclusions: a sophisticated model may appear impressive when compared to a bad baseline, yet show marginal improvement against a well-chosen reference model.

The implications of baseline selection extend beyond individual model evaluation. In collaborative forecasting efforts, such as the COVID-19 and influenza forecast hubs, multiple research teams submit predictions that are evaluated against common baselines. The choice of baseline affects not only which models are considered successful but also how different models are ranked relative to each other. However, the baseline choice can systematically alter model rankings only when forecast models do not fully overlap in their spatial or temporal coverage, a common situation in forecast hubs where different teams may focus on different regions or periods. In such cases, an improperly chosen baseline can introduce systematic biases, making it difficult to identify where and when certain forecasting approaches truly perform well.

Despite its importance, there is no consensus on what constitutes a “good” baseline model. This lack of standardisation is problematic for several reasons. First, it reduces comparability across studies, different baseline choices can lead to conflicting conclusions about the same forecasting method. Second, without clear guidelines, researchers may inadvertently select baselines that favour or disadvantage their proposed methods, either by choosing overly simplistic references or by selecting baselines that perform in a particular way on the specific dataset. Third, the absence of structured baseline selection procedures makes it difficult to determine whether observed improvements stem from genuine methodological advances or from comparison against inadequate benchmarks.

The distinction between a good forecasting model and a good baseline model is crucial yet often overlooked. A good forecasting model aims to optimise predictive performance. In contrast, a baseline need not have exceptional predictive performance, nor should it necessarily perform poorly. Rather, it should be reliable, interpretable and fair across the various conditions under which models are evaluated, and thus reflect a minimum standard that any “good” forecast model should be able to outperform.

This paper addresses these challenges by proposing a structured framework for baseline model selection in epidemiological forecasting. We establish criteria for what constitutes a suitable baseline, provide recommendations for baseline selection and evaluation, and demonstrate the consequences of different baseline choices using data from recent collaborative forecasting hubs. Our analysis reveals that baseline selection impacts model evaluation outcomes and that careful consideration of baseline properties, particularly calibration across different subsets of data, is essential for fair and meaningful forecast assessment.

### 1.1 Previous approaches in infectious disease epidemiology

The choice of baseline model often reflects the underlying characteristics of the epidemic time series, including seasonality, trend patterns, and data availability. We here provide a quick overview of some baseline models used across different infectious diseases and other fields. These diverse approaches to baseline model selection highlight that there is an overlap in baseline models for disease but there is no consensus for a suitable baseline across diseases or fields of applications. Further, there is no consensus on how simplicity in the baseline is balanced with the need to capture essential features of the time series.

#### COVID-19

For COVID-19 forecasting, researchers have predominantly employed simple methods that reflect the rapidly evolving nature of the pandemic. The most common approach uses the most recent observation as point forecast, with various methods for uncertainty quantification. Cramer et al (2022) compared a median forecast using the most recent value against a random walk model with dispersion based on the previous five observations for COVID-19 mortality in the US. Similarly, Meakin et al (2022) used a no-change model from the last observation for hospital admissions in England, while Funk et al (2020) employed a constant model with uncertainty derived from a truncated normal distribution for UK forecasts.

More sophisticated baseline approaches incorporated trend components. Bracher et al (2021b) evaluated three baselines for German COVID-19 data: a constant model, multiplicative extrapolation, and multiplicative smoothing without seasonality. Petropoulos and Makridakis (2020) applied exponential smoothing with trend but no seasonal component for global COVID-19 forecasts, capturing rapid growth phases.

#### Influenza

Influenza forecasting baselines often incorporate seasonal patterns, reflecting the periodicity often observed in influenza epidemics. Reich et al (2019) and Ray and Reich (2018) both employed seasonal kernel density estimation (KDE) based on historical seasons for US influenza forecasting, with Reich et al (2019) also considered the Delphi Uniform baseline that assigns equal probability to all outcomes. Colón-González et al (2021) used seasonal averages for dengue forecasting in Vietnam, citing similar approaches in influenza studies.

Several studies suggested more complex baselines. Kandula and Shaman (2019) argued for non-naïve baselines and employed ARMA-like models for influenza-like illness (ILI) in the US. Morris et al (2023) used both a constant model and a simple elastic net as comparisons for neural network models. Recent work by Ray et al (2025) compared a flat baseline with one incorporating local trends, while Wang et al (2024) evaluated both the most recent observation and historical averages across 32 countries.

#### Other diseases

For Ebola forecasting, Viboud et al (2018) used an AR(3) model on synthetic data informed by the 2014-15 outbreak in Liberia. Funk et al (2019) compared three baselines for Sierra Leone data: a deterministic SEIR model, an unfocused incidence stochastic volatility model, and a Bayesian AR(1) model.

Baseline models for vector-borne diseases often need to account for complex seasonal patterns and environmental drivers. Dengue forecasting has employed various seasonal baselines. Johansson et al (2019) used both a null model with equal probability and a SARIMA baseline for South American dengue forecasts. Lauer et al (2018) utilized a 10-year median for Thailand, while Colón-González et al (2021) applied seasonal averages in Vietnam. For Zika forecasting in Latin America, both McGough et al (2017) and Lewnard et al (2014) employed autoregressive models, with McGough et al (2017) using an AR(3) as a simple baseline and Lewnard et al (2014) considering (S)ARMA models. While many applications include only a single reference model, Keyel and Kilpatrick (2023) recommend inclusion of multiple models.

#### Other Fields

The selection of baseline models in epidemiological forecasting has parallels in other forecasting domains. Gneiting et al (2006) established the no-change forecast as a standard reference in meteorology. In economic forecasting, Lerch et al (2017) used AR(p) models with constant variance for inflation and GDP data, while Krüger (2017) employed the last observation with uncertainty estimated from Gaussian variance fitted to the previous 20 observations for Euro area growth and inflation forecasts. While domain-specific baselines can provide valuable context-specific benchmarks, we focus exclusively on statistical models to develop a general baseline selection framework that can be applied across diseases, outbreak phases, and forecasting contexts without requiring specialised epidemiological knowledge or external data sources.

## 2 Methods

In the context of distributional forecasting, a good forecast model and a good baseline serve different purposes and should be evaluated by different standards. A good forecast model is typically more complex. It aims to predict a distribution that aligns with the truth, usually rewarding performance in accordance with the forecasting paradigm of maximising sharpness subject to calibration (Gneiting et al, 2007). In contrast, a good baseline is generally naïve and simple, designed to provide a stable and interpretable reference point. It should be robust in the sense of reliable performance across different conditions, making it hard to beat globally.

### 2.1 Baseline Models

We restrict ourselves to statistical models that are suitable as a baseline due to their simplicity in terms of computational complexity and interpretability. In total, we include ten statistical model frameworks which can be grouped into naïve models, simple regression models, autoregressive models and time series decomposition models. Despite the spatial correlation we often observe in infectious disease data, we do not account for it in the baseline models. Instead, the models are fit to data for spatial units separately. To introduce the models formally, let *y*_*t*_ be the number of infections observed for a selected spatial unit at time *t* ∈ {1, …, *T*}. Further, we denote 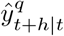 as the forecast of the *q*% quantile for week *t* + *h*, based on the data available up to week *t*. We denote the point forecast as *ŷ*_*t*+*h*|*t*_, often coinciding with the median prediction 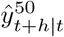. Given a set of probabilities *Q* = (*q*_1_, …, *q*_*k*_)^*′*^, we denote the discretised forecast distribution as 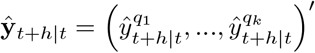 _*′*_.

#### Constant Model

As a very simple naïve baseline model, we use the constant model, also referred to as “random walk”,

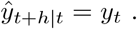

#### Marginal Model

The marginal model is a generalisation of the constant model, where the point forecast is the average of the *p* most recent observations,

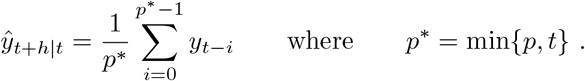

It reduces to the constant model for *p* = 1 and is a true marginal model for *p* = ∞.

#### Kernel Density

A similar alternative to the marginal model is the kernel density model (KDE) that approximates the marginal distribution of a time series. Using a kernel *G* with bandwidth *w*, and training data *y*_1_, …, *y*_*t*_ the marginal density is approximated as

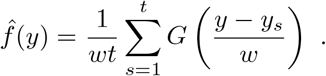

#### Last Similar Date

To account for seasonality in time series data, we include a model (LSD) that is similar to the marginal model but accounts for seasonality. Given an integer periodicity *s*, we may predict that the number of infections is similar to the number of infections at the same week of the year in prior seasons. While being useful in applications with a long history of available data, infectious disease data is often not available for a longer time. Thus, we generalise the idea and include not only the same week of the year, but also similar dates. To this end, let *w* be a non-negative integer. Point forecasts are computed as

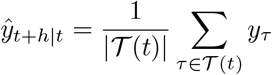

where 𝒯(*t*) = {*t* −*ks*− *w*, …, *t*− *ks* + *w*| *k* = 0, …, ⌈*t/s*⌉}∩ {1,…, *t*} collects all similar dates. The model contains the “last equivalent date” model as special case for *w* = 0.

#### Ordinary Least Squares

Allowing for local trends, we include a simple regression baseline. Given the most recent *p* observations and a polynomial order *d*, we fit an OLS model *y*_*t*_ = *β*^*′*^*x*_*t*_ + *ϵ*_*t*_ with *x*_*t*_ = (1, *t*, …, *t*^*d*^)^*′*^ locally to *y*_*t*_ − *p* + 1, …, *y*_*t*_. The point forecast is

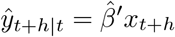

where 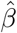 is the OLS estimate.

#### Increase-Decrease-Stable

The Increase-Decrease-Stable model (or IDS for short) fits an OLS model locally and includes a linear trend only if the most recent *p* observations have a clear direction, i.e. *d* = 1 if *y*_*t*_ < *y*_*t−*1_ < … < *y*_*t−p*+1_ or *y*_*t*_ *> y*_*t−*1_ *>* … *> y*_*t−p*+1_ and *d* = 0 otherwise.

#### ARMA

A popular and flexible model framework in forecasting is the ARMA model. We allow the mean of the process to be described by a function *µ*_*t*_(*θ*), parametrised by *θ*. This mean function may be constant, contain a sine-cosine seasonality term or a linear trend. We fit

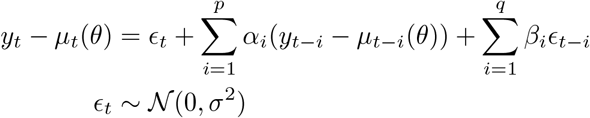

to all observations up to *t*. We estimate the ARMA parameters *α* and *β* together with *θ* by Maximum Likelihood.

#### INARCH

The INARCH model, introduced by Ferland et al (2006), is defined for integer-valued time series. We denote its autoregressive order as *p*. In case seasonality is taken into account, we include sine-cosine waves where *s* denotes the periodicity and *K* gives the number of harmonic waves. Let log(*µ*_*t*_) = *β*^*′*^*z*_*t*_ be the seasonality term where *z*_*t*_ ∈ ℝ^2*K*^ collects sin(2*πkt/s*) and cos(2*πkt/s*) for *k* = 1, …, *K*. We fit

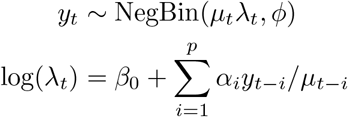

by Maximum Likelihood.

#### Exponential Smoothing

We include exponential smoothing of the target time series with the ETS model, see for example Chapter 7 in Hyndman and Athanasopoulos (2018) and the *forecast* package in R (Hyndman and Khandakar, 2008). The general idea is to decompose the observable time series *y*_*t*_ into error, seasonality and trend.

Each component, if included, can be additive or multiplicative, the trend component can further be damped or not. Therefore, we have two choices for the error, three for the seasonality and five for the trend and thus 30 models in total. The periodicity of the seasonal component needs to be selected. The general framework can be expressed as

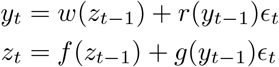

where *z*_*t*_ is a latent vector containing the trend and seasonality component and *ϵ*_*t*_ is the error. The functions *w, r, f* and *g* are determined by the model selection.

#### STL

The season and trend decomposition using LOESS (STL) also decomposes the time series into seasonality, trend and remainder

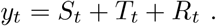

The model is fitted iteratively using a locally weighted regression (LOESS). If a seasonal component is included, only its periodicity must be provided. Tuning coefficients that determine the number of iterations in the estimation and the extent of smoothing are set to reasonable default values. For details, see Cleveland et al (1990).

### 2.2 Evaluation

The *Weighted Interval Score* (WIS) is a proper scoring rule for evaluating predictive distributions, particularly useful when forecasts are given in terms of quantiles or prediction intervals (Bracher et al, 2021a). When evaluating multiple forecast distributions jointly, individual WIS values are aggregated by taking the average, which maintains propriety. If forecasts are evaluated on the log-transformed scale, we denote the WIS as *lWIS* in the following. Applying the score on a log-transformed scale is especially meaningful in epidemiological applications, because it evaluates the predictive performance of the growth rate. Hence, less emphasis is put on times of declining infections after a peak and more on early outbreaks (Bosse et al, 2023).

When evaluating multiple models with forecasts on different spatial or temporal subsets, pairwise relative scores, say 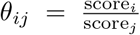 for models *i* and *j*, need to be harmonised. One approach is averaging pairwise relative scores using the geometric mean (Bosse et al, 2022b). Each pairwise comparison is based on the set of overlapping forecasts of models *i* and *j* and are set to one in case of no overlap. The relative skill score of model *i* is the geometric mean 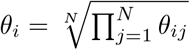, where *N* is the total number of models to be compared including the baseline model. Dividing all relative skill scores by the baseline’s relative skill score gives the scaled relative skill score (SRSS). This approach ensures comparability even when models differed in forecast coverage. A lower SRSS value indicates better forecast performance.

*Forecast calibration* refers to the agreement between predicted probabilities and observed frequencies. A calibrated forecast provides prediction intervals that contain the true outcome at the expected rates over time. In contrast to performance metrics such as the WIS, which evaluate the accuracy forecasts encompassing aspects of both calibration and sharpness, calibration on its own reflects the ability of a forecaster to quantify their own uncertainty, irrespective of how precise the forecasts are. For baseline models, the primary goal is to achieve good calibration, regardless of its precision.

A useful visual tool to assess the calibration of forecasts for integer-valued target variables is the *non-randomised probability integral transform (PIT) histogram*, see Czado et al (2009). Suppose that forecasts are given in terms of *K* central prediction intervals and a median forecast *m*. This yields a set of *M* = 2*K* + 1 quantile forecasts with levels *q*_1_, …, *q*_*M*_. For completeness, we define *q*_0_ = 0 and *q*_*M*+1_ = 1, assigning forecast values of −∞ and ∞, respectively. Given a time series of observations *y*_*t*_, let *l*_*t*_ be the largest forecast value smaller than *y*_*t*_, and *u*_*t*_ the smallest forecast value greater than *l*_*t*_, with corresponding quantile levels 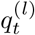 and 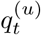.Then, the PIT function for a single observation *y*_*t*_ is defined as

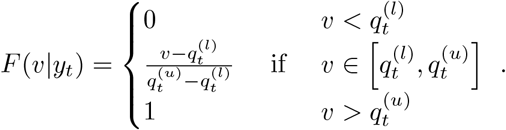

The PIT function for the full time series is obtained by averaging over time:

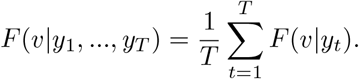

If the forecasts are well calibrated, the PIT function approximates the cumulative distribution function (CDF) of a uniform distribution on [0, 1].

We can quantify deviations from perfect calibration using the *Cramér-von-Mises (CvM) divergence* between the PIT function and the uniform CDF:

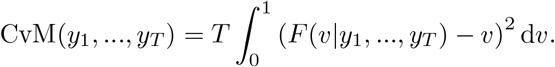

This quantity is easy to compute since the PIT function is piecewise linear between quantile levels. Henceforth, we refer to the CvM divergence as (global) calibration. When assessing calibration on different subsets of forecast data, such as separately for states, we define *uniform calibration* (with respect to a partitioning) as the sum of CvM divergences of subsets.

### 2.3 Recommendation /Guidelines

When constructing a baseline model, it is essential to assess its suitability for evaluating the performance of more complex models. The following steps outline key criteria to consider when selecting a baseline model.

1. **Key Characteristics**. Typical properties of time series, particularly in epidemiological contexts, may include positivity, integer-valued observations, periods of exponential growth, and seasonal effects. A baseline should account for key characteristics if they are undeniable.
2. **Format Compatibility**. Forecast performance metrics require comparability of outputs. If the baseline does not generate forecasts in the same format (e.g. quantiles, predictive distributions), fair evaluation is not feasible.
3. **Training Data Availability**. The baseline must not have access to more or better training information than the evaluated model. This usually includes using only the type of data that is being forecast for training and excluding any other potentially informative data sets.
4. **Simplicity**. A good baseline should be easy to fit, interpret, and communicate. Overly complex baselines defeat the purpose of providing a reference.
5. **Calibration**. A good baseline should be calibrated globally, i.e. across all spatial units, time points and forecast horizons. Ideally, a baseline model is also well-calibrated in different strata.

If a baseline does not account for key characteristics of the data, it may overly favour the forecast model. Conversely, if the baseline makes implausible assumptions about these characteristics, it may lack calibration, leading to poor forecast performance. A baseline that is not well-calibrated overall can distort the evaluation. More importantly, if the baseline is not well-calibrated in different strata, the performance of a single forecast model is not comparable between these strata in a fair way.

These recommendations can be systematically evaluated using diagnostic procedures. To assess the appropriateness of log-transformation, one can apply the Box-Cox transformation and examine whether the estimated power parameter is near zero. For multiple time series, histograms of power parameters can be used to assess whether consistent transformation across the dataset is reasonable. The approach extends to other transformations, such as a square root transformation. Time series characteristics can be tested visually using the autocorrelation function at lag one and suspected seasonal periods. Through ordinary least squares regression of the (appropriately transformed) series on lagged observations, sine-cosine seasonal terms, and linear time trends, one can test these characteristics with standard t- and F-tests. Global calibration can be evaluated visually through PIT histograms and quantified using the Cramér-von-Mises divergence. Deviations from perfect (global) calibration can be tested with bootstrap tests sampling quantile bins according to their nominal probability levels to assess deviations from perfect calibration. For uniform calibration, PIT functions stratified by location, time, or forecast horizon can reveal systematic biases across subsets, and bootstrap procedures can test both for deviations from perfect uniform calibration and for differences between strata, ensuring fair model comparisons across all evaluation contexts. However, formal testing is not always necessary. Domain knowledge often provides guidance about expected time series characteristics, such as known seasonal patterns in influenza or obvious autocorrelation in epidemic data, allowing researchers to proceed with theoretically motivated baseline specifications.

## 3 Data

This study utilises publicly available forecast and ground truth data for both seasonal influenza and COVID-19, focusing on weekly hospitalisation counts in the United States. All forecasts from forecast models and baseline models for influenza consist of a forecast distribution stored as a set of quantiles for probabilities 5%, 10%, …, 95% as well as 1%, 2.5%, 97.5% and 99%. For COVID-19, only seven quantiles are available (2.5%, 10%, 25%, 50%, 75%, 90%, and 97.5%). We combine the forecast distributions made at the same reference date, for one location across different horizons as one forecast.

### 3.1 Forecast Data

#### 3.1.1 Influenza

Influenza forecasts were sourced from the CDC FluSight initiative, including both archived and the current repository, see CDC Influenza Forecasting Team (2021) and CDC Influenza Forecasting Team (2025). Forecasts span three complete influenza seasons (2021/22 through 2023/24) and one incomplete season (2024/25) and include submissions from 86 unique models across 110 forecast dates, covering all 50 U.S. states, Washington D.C., and national-level data. Forecast data from both repositories were harmonised through the following steps:

Only quantile-based forecasts were retained (23 quantiles per forecast). Forecasts were filtered to include only the target variable: weekly incident hospitalisations. Forecasts covering U.S. territories and county-level data were excluded. Only future-facing horizons were kept for analysis. For the newer repository, horizons −1 and 0 (representing retrospective or current-week forecasts) were excluded. The cleaned data were merged into a unified format with metadata including model, forecast date, location, target, horizon, and quantile. A summary of the resulting dataset is provided in the supporting information. After cleaning, we were left with 81 models. Out of all models, only six included one location, one model only covers six locations, seven models submitted forecasts for all states and not the national level. With 67 models, the majority submitted forecasts for all locations considered in this study. For season 2021/22 and 2022/23 forecasts are available from around 30 models each. In seasons 2023/24 and 2024/25, we are left with forecasts of around 40 models each.

#### 3.1.2 COVID-19

COVID-19 hospitalisation forecasts were obtained from the COVID-19 Forecast Hub Reich Lab (2025a). The initial dataset comprised 129 models with submitted forecasts for a variety of targets (e.g., cumulative deaths, incident hospitalisations). For this study, only quantile-based forecasts (7 quantiles) of weekly hospitalisations at the state and national levels were retained. The filtering process involved exclusion of forecasts for other outcomes (e.g., deaths), daily predictions, and county-level data, as well as of models that did not provide the required forecast structures (quantiles over relevant horizons). The final set included 57 models with hospitalisation forecasts across 152 weeks and 52 locations. For eight models, there were only forecasts for one location available. For the remaining models, forecasts had been submitted for most locations, with nine models where the number of locations is slightly below 52 and 40 models that submitted forecasts for all locations. On average, forecasts were available for 67 weeks. The CovidHub baseline model was the only model with full coverage of weeks and locations. A table of available forecasts is provided in the supporting information.

### 3.2 Truth Data

Ground truth data for hospitalisations served both as evaluation basis and training inputs. For influenza, these data were included within the forecast repositories. We discarded the initial 40 weeks with no cases nationwide. For COVID-19, truth data were obtained using the *covidHubUtils* R package, which provides access to hospitaliation data from publicly available sources (Reich Lab, 2025b).

## 4 Application

We followed a similar process for both influenza and COVID-19 data. First, we selected an appropriate model architecture for each baseline model framework. To evaluate the baseline models, we discuss their suitability qualitatively as well as quantitatively using calibration metrics. Finally, we examined how the choice of a baseline model influences the evaluation of forecasts submitted to the forecast hubs.

### 4.1 Baseline Settings

A defining feature of influenza time series is their strong seasonal pattern, which, when relevant, should be captured by a baseline model. We assumed temporal autocorrelation in the data.

For the constant model, no architectural choices were required. The marginal model uses all available observations. The kernel density model was applied with default settings. The least-similar-date model assumed a 52 week periodicity and used a window of eleven observations, i.e. weeks were considered similar if they differed by no more than five weeks. This approach balances the trade-off between data availability and temporal similarity.

OLS models were fitted using the five most recent observations and included a linear time trend to capture short-term upward or downward movements in infection counts. The IDS model relied on the three most recent observations. For the ARMA model, we specified orders *p* = *q* = 1 and assumed a 52 week periodicity and no time trend in the marginal mean function. The INARCH model was fitted with order *p* = 1 incorporating a 52 week seasonal pattern represented by *k* = 4 harmonic waves. The conditional distribution was modeled as Negative Binomial.

The ETS model was applied with additive errors, no trend component, and used pre-filtered seasonality: an additive seasonal component with 52 weeks periodicity was estimated and removed prior to fitting, and then added back to the forecasts. Similarly, the STL model was applied with default settings, and the seasonal component was handled through the same pre-filtering and post-correction process.

Unlike influenza, COVID-19 time series have not exhibited a regular seasonal pattern but often feature phases of exponential growth. As in the influenza case, the constant model required no architectural choices and the marginal model utilises all past observations. The kernel density model was again applied using default settings. The LSD model - due to the absence of seasonality - reduced to a marginal model. A window size of eleven translates to a marginal model based on the five most recent observations.

The OLS and IDS models were specified the same as for influenza. For the ARMA model, we specified orders *p* = *q* = 1, no seasonality, but include a linear time trend in the marginal mean function. The INARCH model was used with order *p* = 1, no seasonality, and a Negative Binomial conditional distribution.

For the ETS model, we used multiplicative errors, a damped additive trend, and no seasonality. The STL model was applied with default settings and excluded a seasonal component.

To account for positivity of the time series, we included all models except for the INARCH model to also be applied to log-transformed data. For the ETS model and COVID-19, we substituted multiplicative errors with additive errors and left all other settings the same as for an application to non-transformed time series.

Baseline model fitting is performed in Julia using the ForecastBaselines.jl package (Stapper, 2025). The package offers three different ways to compute prediction intervals: Based on historic forecast errors (*EmpiricalInterval*), by running chains of model trajectories into the future (*ModelTrajectory*) or analytically (*Parametric*). The latter two depend on the model chosen; for details, see the package documentation.

The empirical interval method is based only on historic forecast errors, making every model compatible for which point forecasts can be computed. Let 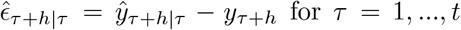 be a historic forecast error. We collect the errors as 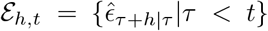. If we want to ensure that forecast intervals are centred around the point forecast, we can include forecast errors of both signs 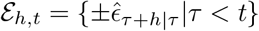. Trajectories are sampled into the future directly from the set of forecast errors instead of fitting a distribution to the forecast errors. We sample h-step ahead trajectories from historic h-step ahead forecast errors instead of successively sampling from one-step ahead forecast errors. To ensure positivity of all forecast quantiles, there are three options: The sampling distribution in each step can be truncated or censored to have positive support or negative forecast quantiles can be censored to zero after sampling is completed. We used this method for the constant, marginal and LSD model. For non-transformed data, we truncate the sampling distribution at zero and did not include forecast errors of both signs to account for asymmetry in the distribution of infections. For log-transformed time series, no correction for positivity was necessary and forecast errors of both signs were considered.

The model trajectory method is used for regression-based baselines OLS and IDS, the INARCH model and for decomposition models ETS and STL. The sampling distribution was truncated at zero for non-transformed data. The KDE and ARMA baselines used the parametric prediction interval method and clipped intervals at zero afterwards.

### 4.2 Baseline Evaluation

The time series of infection counts have fundamental properties that are ideally captured by baseline models. The positivity of time series was accounted for by selecting appropriate prediction interval methods or by applying a log-transformation.

For influenza time series, the constant, marginal, KDE, OLS, and IDS models did not explicitly account for seasonal patterns. In contrast, the ARMA, INARCH, ETS, and STL models incorporated seasonality in their structure. Among all baseline models, only the INARCH model was specifically designed to handle integer-valued time series.

For COVID-19 time series, which often exhibit phases of exponential growth or decline, the OLS, IDS, and STL models could capture local linear trends. The ARMA and ETS models instead reflected broader, overall trends and might not adapt well to sharp changes. Short-term temporal correlation was modeled by the constant, LSD, OLS, IDS, ARMA, INARCH, ETS, and STL models, each incorporating some form of dependency on recent observations.

All models produced forecasts in a format compatible with the forecast hub data. They relied only on past observations from the individual time series for training. However, forecasts from forecast hubs are based on time series available at that time. Retrospective corrections of reported numbers are not accounted for by forecast models. Baseline models are based on retrospectively cleaned data and thus using additional information.

We assessed calibration through the PIT function and the resulting CvM divergence. For a single baseline model, all forecasts across points in time, regions and forecast horizons were jointly evaluated. Figure 1 shows the PIT functions of all baseline models for influenza and COVID-19. Highlighted are the baseline models that were calibrated best and worst. In case of influenza infections, we see that the marginal model (applied to log-transformed time series) exhibited poorest calibration. The ETS model (applied to log-transformed time series) had the best calibration among all baselines.

**Fig. 1:**
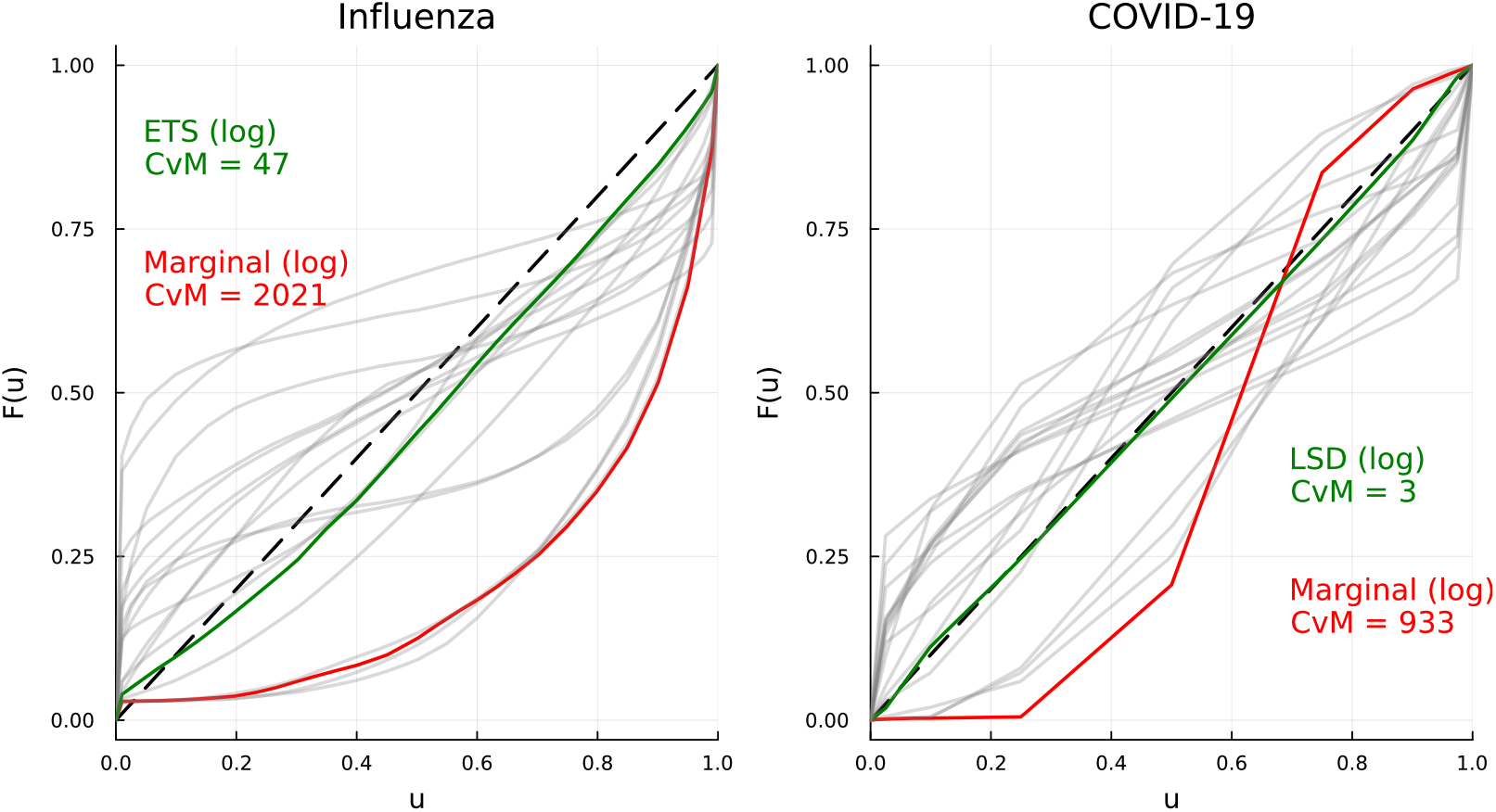
PIT functions for different baselines

The best calibrated baseline for COVID-19 time series was the LSD model applied to log-transformed time series, which computed point forecasts as average of the most recent five observations. The worst calibrated baseline was the marginal model on log-transformed data. This reflects that individual time series had a short-term dependence that needed to be captured for good calibration.

When comparing the performance of a forecast model between subsets of the data, such as for a different horizons, regions or phases, it is important to assess the calibration of the baseline model for the subsets individually. Otherwise, we would risk an unfair assessment caused by differences in the calibration. Figure 2 shows PIT functions for different subsets, split by horizon and region, for influenza and COVID-19 and the corresponding baseline model that was best calibrated on the complete set of forecasts. When split by horizon, the respective best calibrated baseline models showed small differences in their calibration. When looking at PIT functions for different regions, we see that there were larger deviations from the uniform CDF for influenza, whereas for COVID-19 the PIT functions appeared more closely scattered around it.

**Fig. 2:**
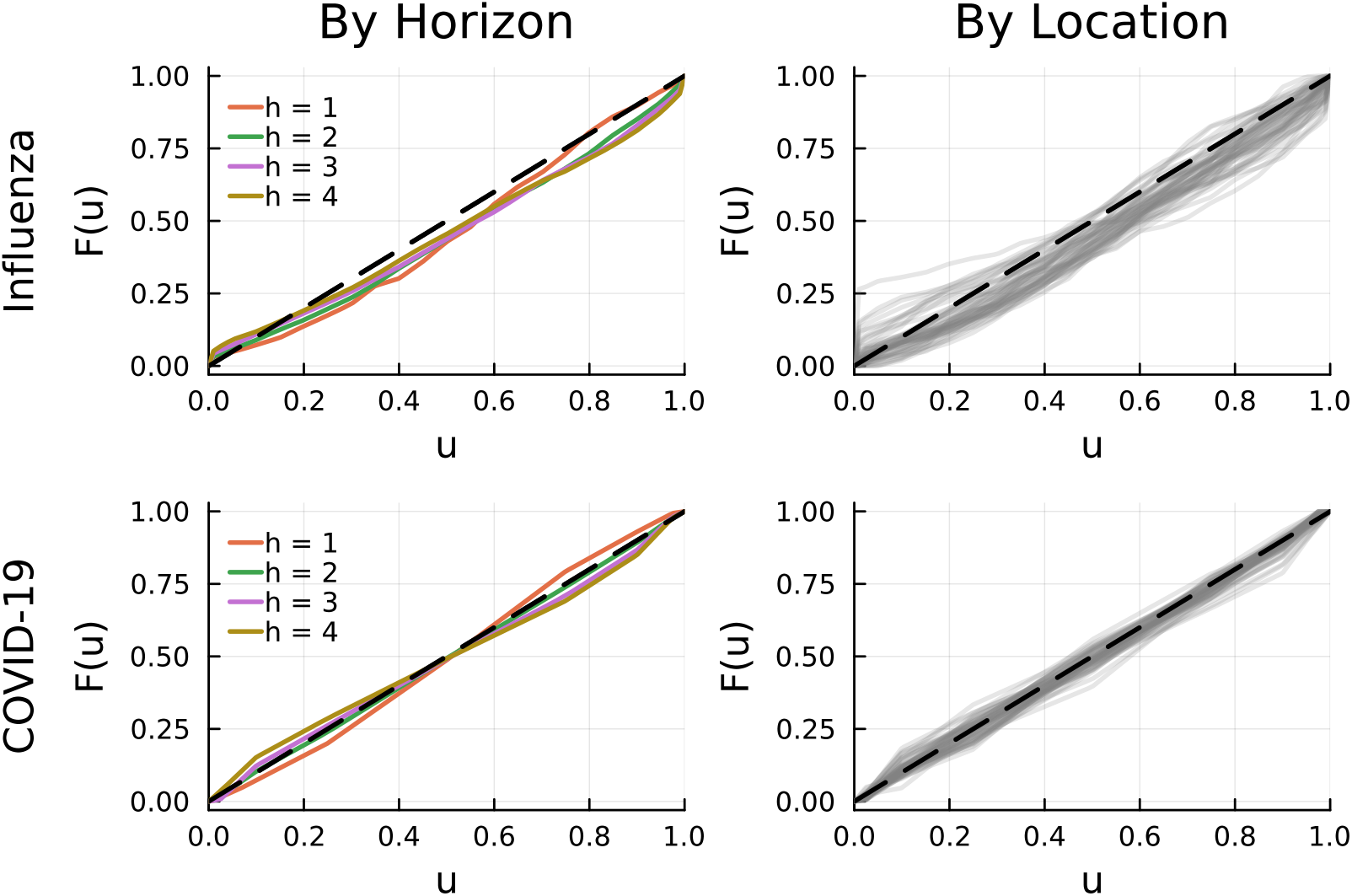
PIT functions for best calibrated baseline - Split by location and horizon.

### 4.3 Baseline Correction

When calibration is not ideal across regions, time periods, or forecast horizons, several strategies can be applied to improve it. One option is retrospective re-calibration of baseline forecasts. By appropriately scaling and shifting the quantile forecasts of a given baseline model, calibration can be improved within specific subsets of the data, potentially improving overall calibration. Another straightforward approach is to allow for different baseline model selections across subsets, such as regions.

Figure 3 illustrates this idea, showing that the best calibrated baseline model differed across regions. For COVID-19, the LSD model (applied to log-transformed data) achieved the best calibration in most regions. In contrast, for influenza, the optimal baseline varied considerably. Across the 52 regions, one of six baseline models was the best calibrated in at least one case. Notably, although the ETS model (in logs) was the best calibrated overall, it was the best calibrated model in only 14 of the 52 regions.

**Fig. 3:**
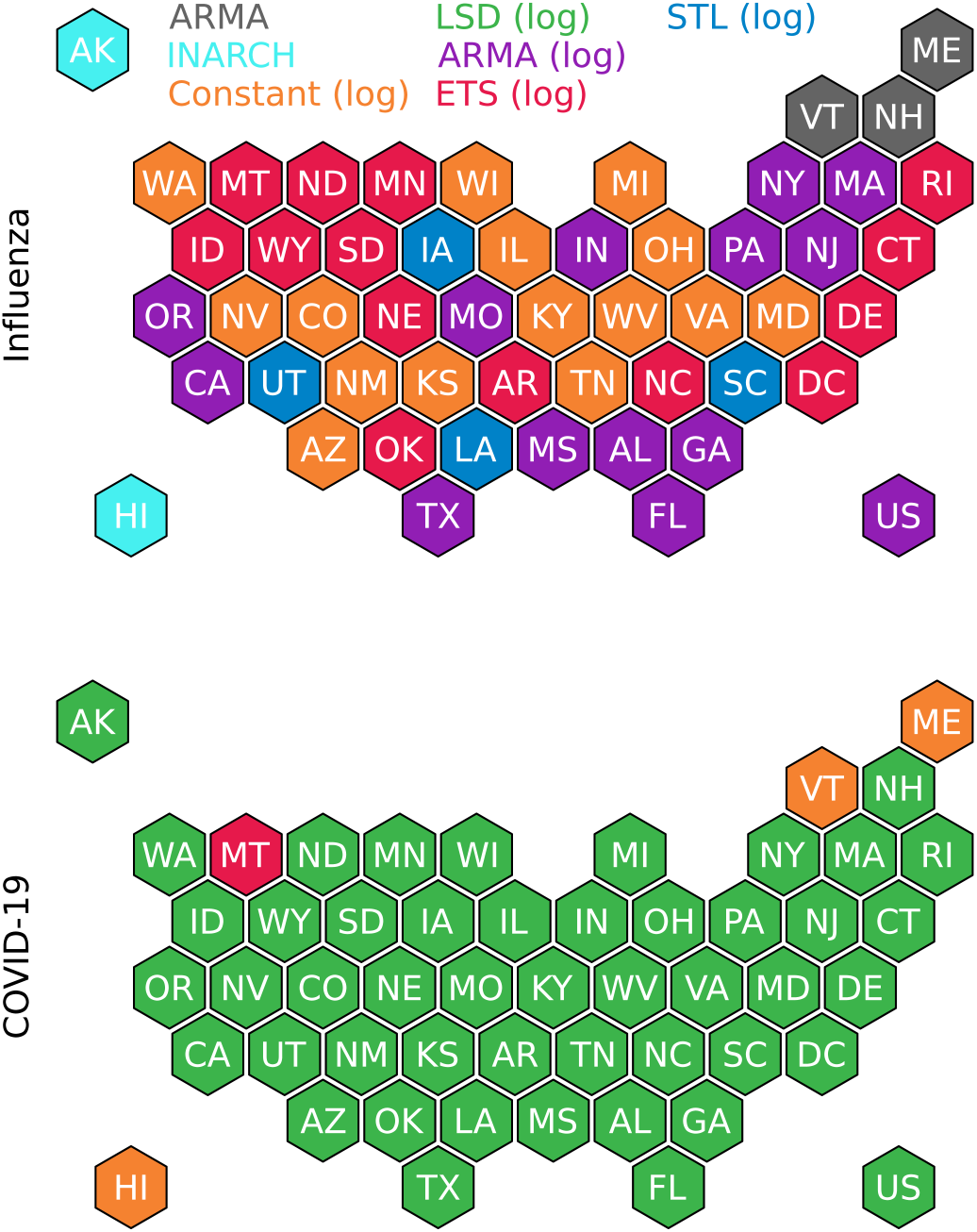
Best baseline model for each US state for influenza (top) and COVID-19 (bottom)

Allowing for region-specific baseline model selection and then aggregating the resulting forecasts can lead to improved overall calibration. For influenza, the CvM divergence for the best single baseline model was around 47 and was reduced to 15 by selecting best individual baselines. For COVID-19 the effect was not present with both, the best baseline and best individual baselines achieving a CvM divergence of around three. The moderate effect is not surprising, since the single best baseline was the best calibrated baseline in most states.

Another strategy for achieving well-calibrated forecasts is based on the following idea: if all observations in a time series for a given region were drawn from the same marginal distribution, and if this distribution was known, we could generate perfectly calibrated forecasts by simply using its quantiles. The marginal and KDE models are motivated by this principle. However, in practice they struggle with calibration because the marginal distribution must be estimated from past data, and the magnitude of epidemiological time series levels can vary over time.

To ensure fair comparison with submitted forecasts, Property 3 of the evaluation recommendations restricted us from using the full data when generating baseline forecasts. Nonetheless, for illustrative purposes, we included an additional marginal distribution baseline in the analysis. The baseline computes quantile forecasts as empirical quantiles of the time series, excluding the target observation and every observation for which there is no forecast available from forecast hubs.

Both additional baseline models were included to achieve better uniform calibration, i.e. improved calibration across subsets of the forecast data. Our focus was on calibration for all states. A baseline that demonstrated good calibration across all states would enable fair comparison of a single model’s forecast performance between states. We assessed this by computing PIT functions separately for each state. Figure 4 presents PIT functions for the two additional baseline models alongside the baseline exhibiting the best global calibration. The PIT functions were summarised as median values with pointwise 80% intervals and ranges across all states. While a baseline with good global calibration would exhibit a median PIT close to the uniform CDF, a baseline with good uniform calibration would additionally display narrow intervals around it. For both influenza and COVID-19, the best single baselines exhibited wider intervals compared to the two additional baselines. Selecting different baselines for individual states did not substantially improve uniform calibration in either case. However, the marginal model that utilises future data significantly improved uniform calibration. For influenza, despite showing near-perfect global calibration, there were deviations from perfect uniform calibration, particularly for small quantiles. These deviations are likely attributable to the large number of zero observations. For COVID-19, we observed near-perfect global and uniform calibration.

**Fig. 4:**
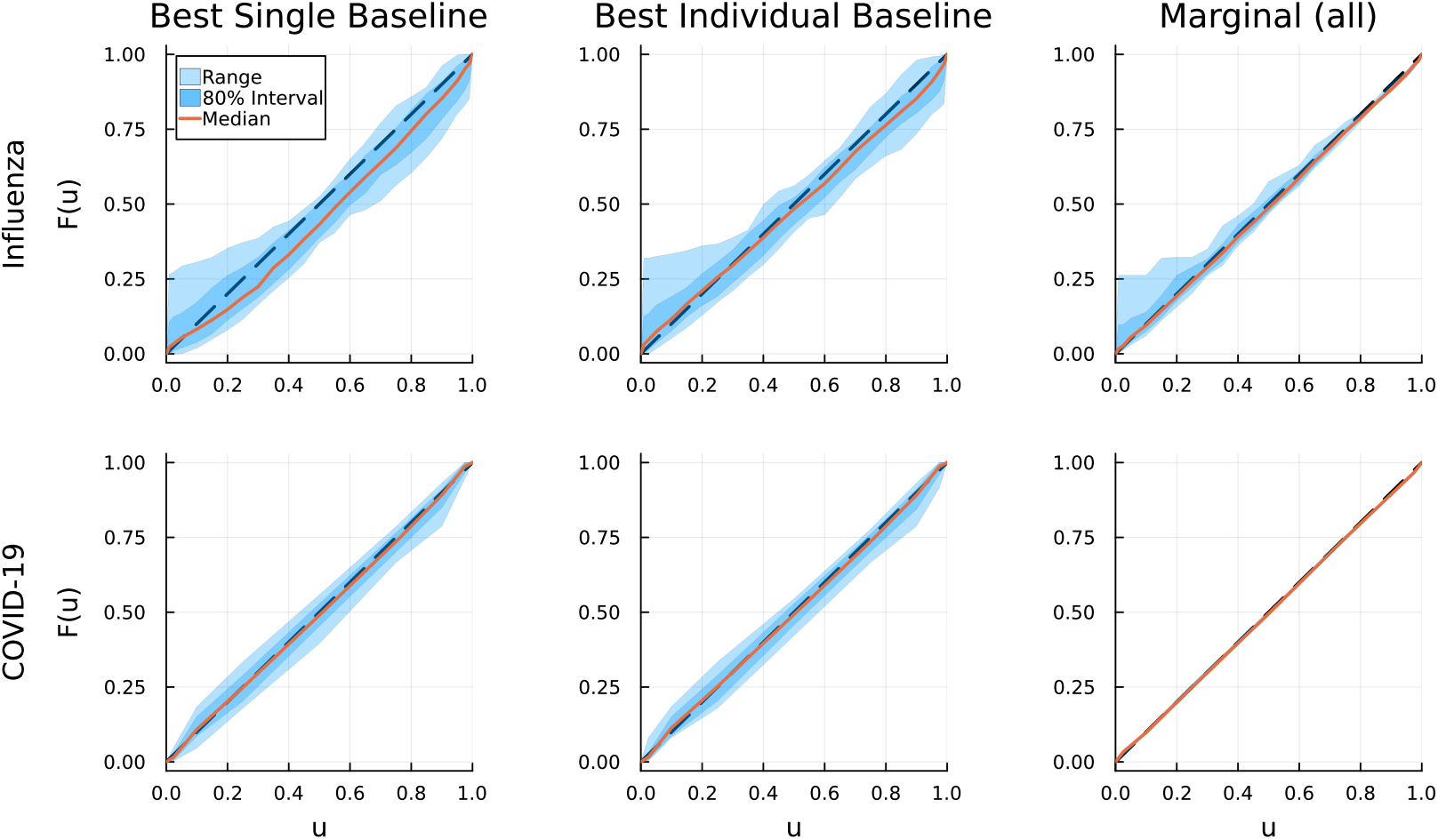
Uniform calibration plot: Pointwise 80% interval (dark blue), range (light blue) and median (orange) of PIT functions across all states

### 4.4 Forecast Evaluation

When comparing baseline forecasts with forecasts from hubs, our objective was to answer three questions:

1. How does the choice of a baseline and scoring rule affect the overall assessment of forecast models?
2. How does the choice of a scoring rule and baseline affect the ranking of forecast models?
3. When considering a single forecast, how does the choice of a baseline affects the evaluation of the model on different subsets, such as regions?

In total, we evaluated forecasts from 81 models for influenza and 57 models for COVID-19 using the scaled relative skill score. For each baseline model, we added it to the pool of forecasting models and computed the scaled relative skill score once based on the WIS on the original scale and once applied to log-transformed forecasts. This gives a total of 38 ranked list of forecasting models. If all baseline models were perfectly calibrated but varied in sharpness, we would expect the overall level in relative skills to vary across the lists, but the ranking to be similar.

Figure 5 shows histograms of SRSS values across forecasting models, split by disease and scoring rule. The comparison focused on models evaluated against either the best or worst calibrated baseline. For influenza, the results show that baseline selection strongly influenced how many models outperformed the baseline. Using the WIS, 9 out of 81 models outperformed the best calibrated baseline, while 75 models out-performed the worst calibrated one. When using the lWIS 9 models outperformed the best-calibrated baseline, whereas 79 beat the worst calibrated baseline. For COVID-19 under the WIS, 56 models outperformed the best calibrated baseline and all 57 models outperformed the worst calibrated one. When using the lWIS, 30 models outperformed the best calibrated baseline and 52 outperformed the worst calibrated one.

**Fig. 5:**
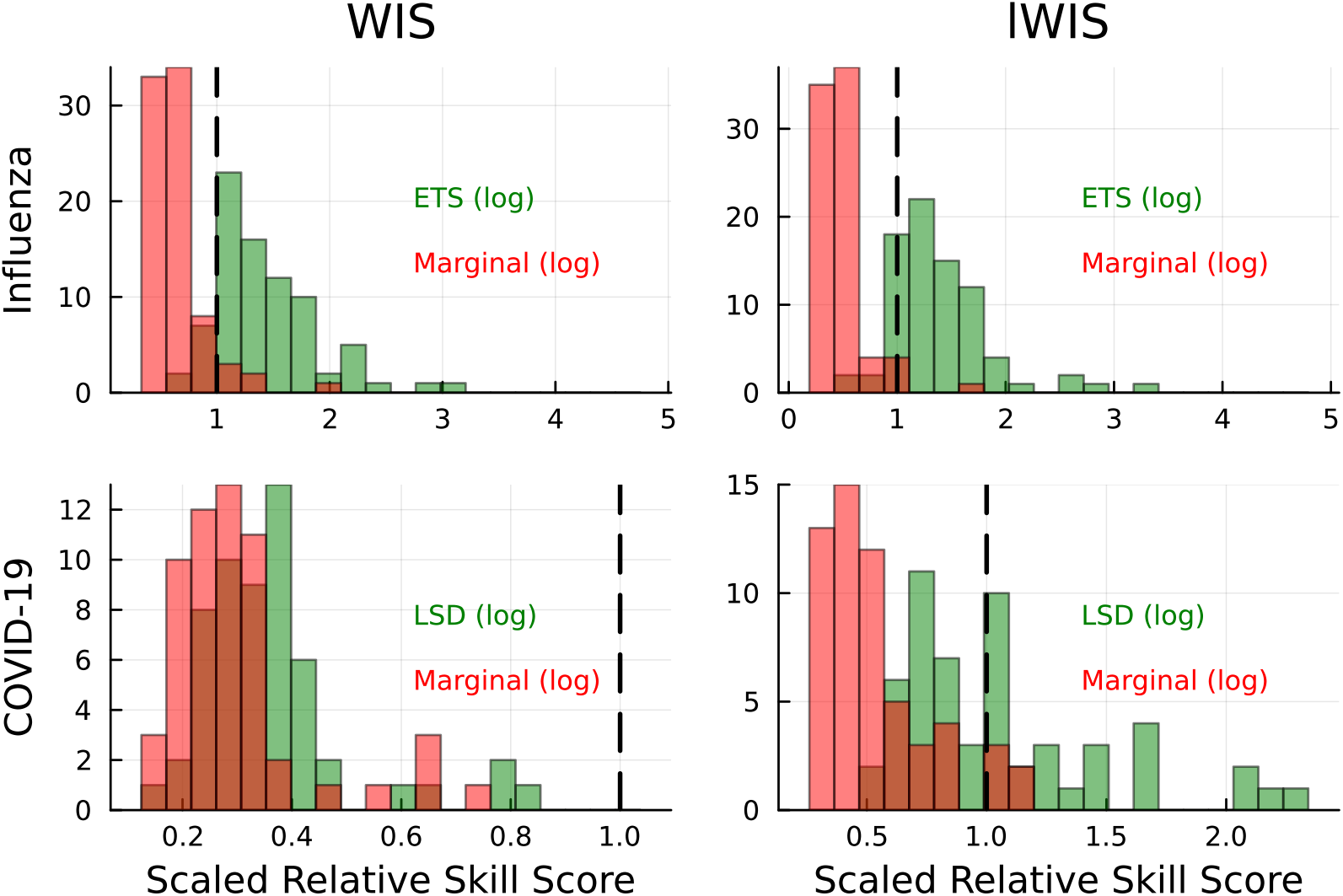
Scaled Relative Skill Scores for best (green) and worst (red) calibrated baselines - Split by disease and evaluation metric

The choice of baseline model can influence not only the overall assessment of forecast models but also their relative performance rankings. Figure 6 displays how forecast models rank under different baseline choices, ordered by the baselines’ calibration quality. If all baselines were perfectly calibrated, we would expect only minor differences between rankings.

**Fig. 6:**
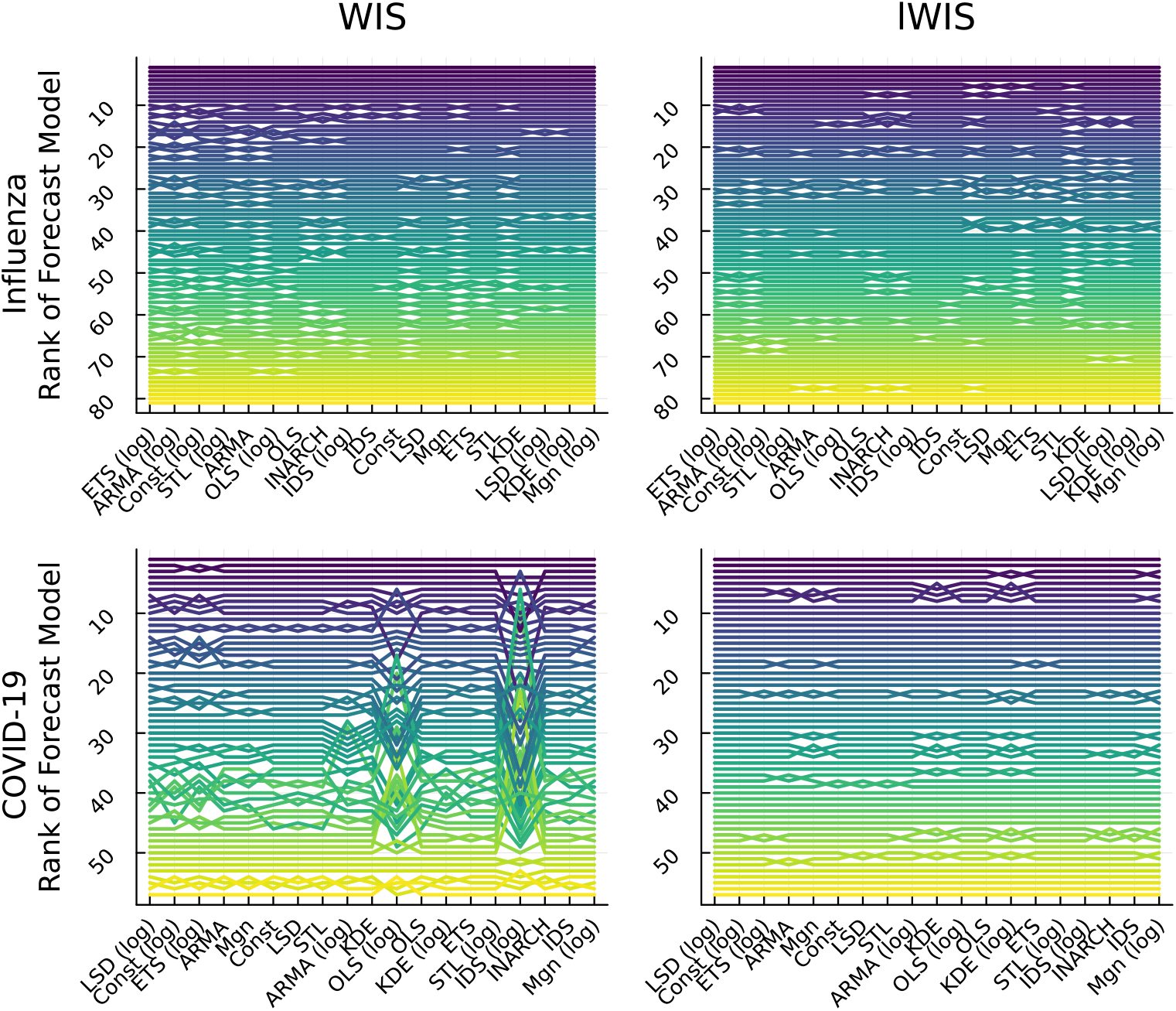
Ranks of forecast models for different baselines - each line represents one forecast model

For influenza, the best and worst performing forecast models maintained their positions regardless of the baseline choice for both scoring rules. For COVID-19 under the WIS, two baseline models—OLS-l and IDS-l—produced markedly different rankings. These baselines ranked 11th and 16th respectively in terms of global calibration. However, under the IWIS, forecast model rankings remained consistent across all baselines.

Figure 7 summarises these ranking differences by Kendall’s tau for each pair of rank lists. Correlations were capped at 0.9 for clearer visualisation. For COVID-19 under the WIS, there was less agreement in rankings compared to influenza. Again, the OLS-l and IDS-l baselines showed the least agreement with others. Notably, the two best calibrated baselines, the LSD and constant models in logs, produced different rankings. When using the lWIS as the scoring rule, rankings showed greater consistency across baseline models for both influenza and COVID-19. Therefore, while baseline choice can affect forecast model rankings, the scoring rule affects the ranking and using the lWIS makes rankings more robust across baselines.

**Fig. 7:**
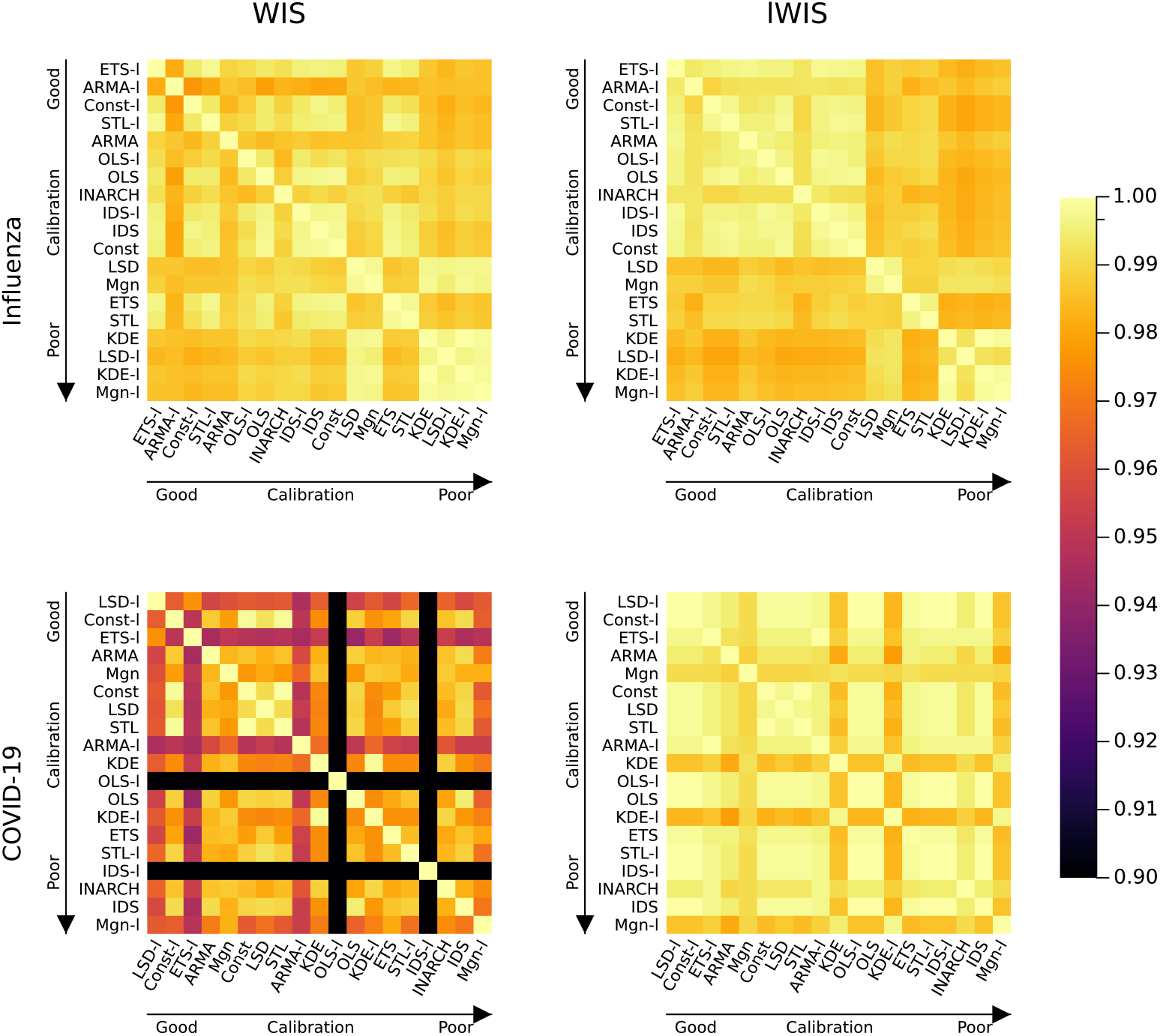
Kendall’s tau correlation heatmap for influenza and COVID-19 based on WIS and lWIS - Correlations are truncated at 0.9

We compared the forecast performance of a single forecast model across individual states. For this analysis, we selected the FluSight-Ensemble and the COVIDhub-Ensemble for influenza and COVID-19 respectively, as these models have good coverage across states and time. We aimed to answer whether the choice of baseline affects our evaluation of where these forecast models perform well. Beyond good global calibration, we assumed that good uniform calibration with respect to state partitioning is necessary to address this question. Figure 8 summarises the uniform calibration of all baseline models, for influenza and COVID-19, alongside their global calibration. The two additional baselines using region-wise best baseline models (“Indiv”) and using all available data (“Mgn-all”) are included for reference. Overall, baseline models with good global calibration were also calibrated well uniformly.

**Fig. 8:**
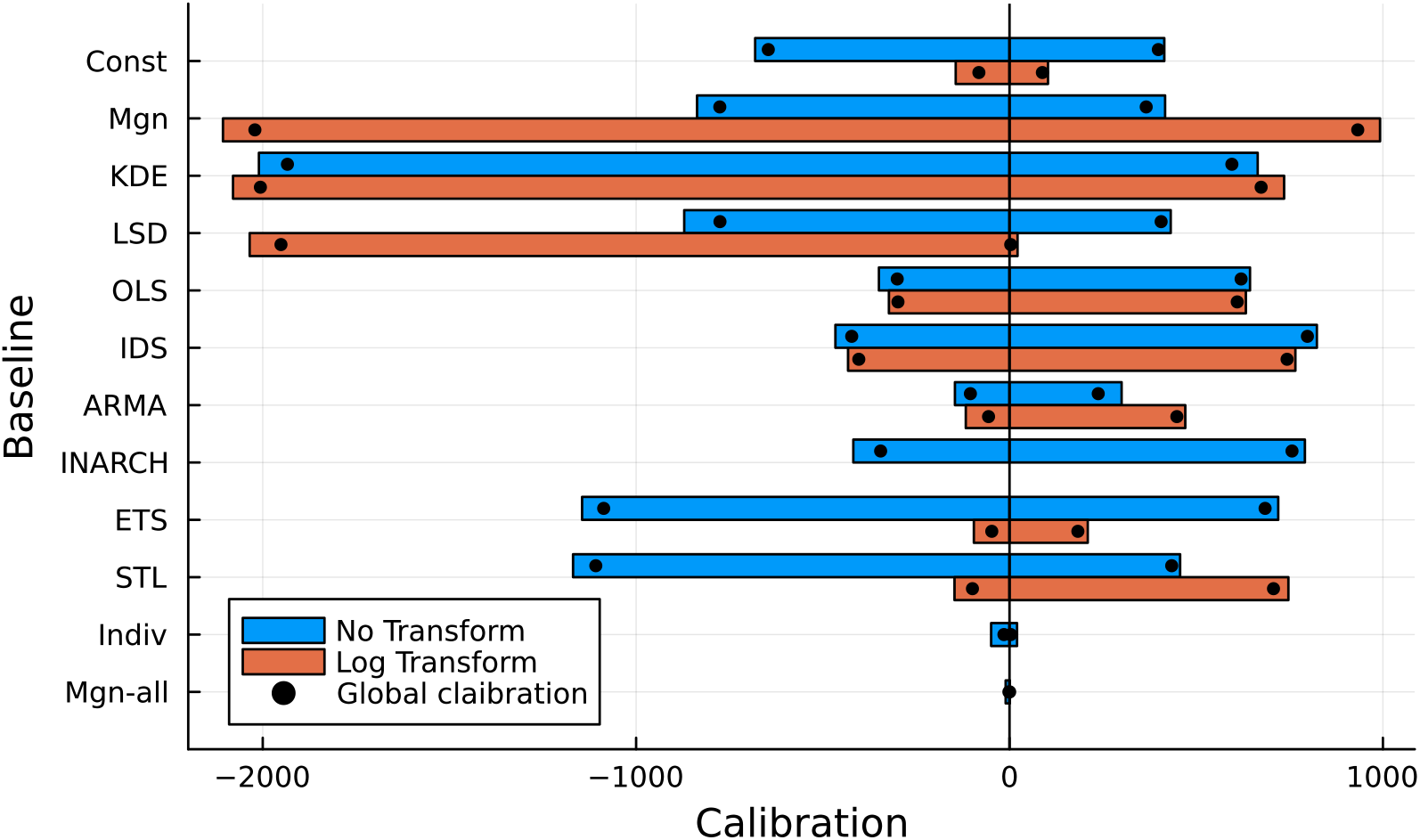
Global and uniform calibration of baseline models for influenza (left) and COVID-19 (right)

We compared the forecasts of the two forecasting models with each baseline’s forecast using the relative WIS and lWIS. Computing the scaled relative skill score was not necessary due to equal coverage across states and time. The relative WIS and lWIS were then used to rank states by the forecast model performance. These ranks were again compared using Kendall’s tau correlation matrices. Figure 9 summarises all correlation pairs, with baselines sorted by their uniform calibration. Negative correlations were truncated at zero. For influenza, there was consensus among the four best (uniformly) calibrated baselines, with strong agreement between the two additional baselines for both scoring rules. For baselines with poorer calibration, there was less agreement amongst themselves and with well-calibrated baselines. For COVID-19, the two additional baselines were again the best calibrated models. Although showing good uniform calibration, the ranking differed between them. Compared to them, many of the other baseline models, regardless of their calibration, have similar correlation in their ranking, suggesting that calibration does not affect the ranking much for COVID-19.

**Fig. 9:**
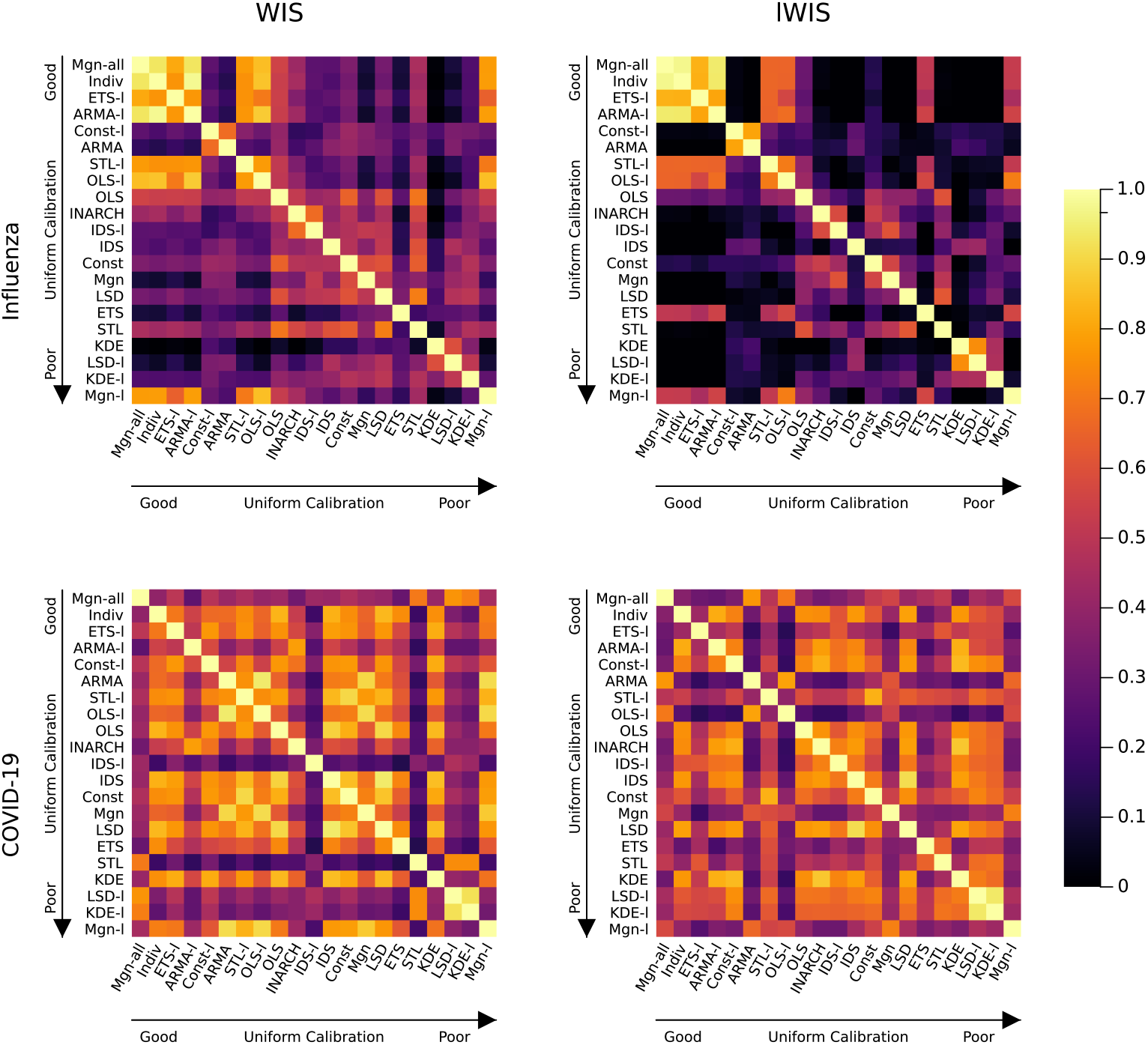
Kendall’s tau correlation heatmap for location ranking - evaluated for FluSight-Ensemble (influenza) and CH-Ensemble (COVID-19). Correlations were truncated at zero.

## 5 Discussion

This study provides recommendations for baseline model selection in epidemiological forecasting, addressing a critical but often overlooked aspect of forecast evaluation. Our analysis revealed that baseline choice profoundly influences both the assessment of individual forecast models and their relative rankings.

Our findings demonstrate that selecting an appropriate baseline model is far from trivial. Among the ten baseline frameworks evaluated, no single model satisfied all recommended criteria across both influenza and COVID-19 applications. The best globally calibrated baseline for influenza differed from that for COVID-19, highlighting that baseline selection is application-specific. We found that calibration and forecast performance do not necessarily align, a well-calibrated baseline may perform poorly in terms of forecast accuracy, while a poorly calibrated baseline may inadvertently favour certain forecasting models.

The impact of baseline selection extends beyond simple performance metrics. When evaluating multiple forecasting models, the choice of a baseline model can alter conclusions about which models are successful. For influenza, the number of models outperforming the baseline ranged from 9 to 75 depending on the baseline selected.

Our analysis of uniform calibration revealed another layer of complexity. A baseline that appears well-calibrated globally may exhibit poor calibration across specific subsets such as individual states or forecast horizons. In our application, however, global and uniform calibration was similar. Heterogeneity in calibration can lead to biased assessments when comparing model performance across different regions or time periods. Such biases have direct practical consequences: if a baseline systematically underperforms in certain regions, forecast models in those areas may appear artificially superior. This potentially leads to resource allocation for prevention and preparedness that is based on a false impression of certainty in forecasts. The finding that regions had different best calibrated baseline models, especially for influenza, suggests that a one-size-fits-all approach may be inadequate for fair evaluation.

From a practical standpoint, we recommend that researchers follow a structured process when selecting baseline models. First, identify the key characteristics of the time series being forecast, including seasonality, growth patterns, and data availability. Second, ensure format compatibility between the baseline and forecast models. Third, verify that the baseline uses appropriate training data. Fourth, prioritise simplicity to maintain interpretability. Finally, and most importantly, assess both global and uniform calibration across relevant data partitions.

When a single well-calibrated baseline cannot be identified, several strategies can improve evaluation fairness. Regional or temporal stratification of baselines may provide better uniform calibration, though at the cost of increased complexity. Alternatively, researchers might consider using multiple baselines to assess the robustness of their conclusions, or employ ensemble baselines that combine simple models. The choice of scoring rule also matters; our results suggest that log-scale evaluation (lWIS) often produces more stable rankings across different baselines than evaluation on the original scale.

This work has several limitations that suggest directions for future research. First, our analysis focused on epidemiological time series, and the generalisability to other forecasting domains remains unclear. Different fields may have distinct requirements for baseline models based on their specific data characteristics and evaluation needs. Second, we restricted our analysis to relatively simple baseline models. While this aligns with the principle that baselines should be interpretable and easy to implement, more complex baselines might achieve better calibration in some settings.

Future research could explore automated procedures for baseline selection, potentially using cross-validation or other model selection techniques to identify baselines that achieve good calibration. The development of diagnostic tools to assess baseline suitability before conducting forecast evaluations would be valuable. Furthermore, the relationship between baseline selection and the predictability of time series is an interesting direction. The forecast performance of a suitable and simple baseline gives insight into how much of the variation in a time series can be explained by a simple model.

The challenge of baseline selection reflects broader issues in forecast evaluation. Just as human judgment serves as an informal baseline in many decision-making contexts, statistical baselines provide a reference point for assessing the value of computational forecasting methods. Our results suggest that this reference point must be chosen carefully, with attention to both statistical properties and the specific evaluation context. At the same time, systematic collection of human judgement forecasts could enable their use as an additional baseline (Bosse et al, 2022a, 2024). As collaborative forecasting efforts continue to grow, establishing standards for baseline selection will become increasingly important for ensuring fair, meaningful, and reproducible model comparisons.

In conclusion, baseline model selection deserves the same careful consideration given to forecast model development. By providing tools to assess baseline calibration and demonstrating the consequences of different choices, this work aims to improve the rigour and fairness of forecast evaluation in epidemiology and beyond. The lack of a universally optimal baseline underscores the need for thoughtful, context-specific selection guided by clear principles and thorough evaluation.

## Data Availability

All data is publicly available and summarised at \url{https://doi.org/10.5281/zenodo.16992035}. Application code is available at \url{https://github.com/ManuelStapper/Baselines_Application}.

https://doi.org/10.5281/zenodo.16992035

https://github.com/ManuelStapper/Baselines_Application

## Acknowledgments

The authors would like to thank the *epiforecast* team members for their valuable contributions to the literature review and project conceptualisation in the early stages that formed the foundation of this research.

## Use of AI

The authors acknowledge the use of Claude Sonnet 4 (Anthropic) for formatting and proof reading. All AI-generated content was thoroughly reviewed, fact-checked, and revised by the authors. The final analysis, conclusions, and academic interpretations are entirely the authors’ own work.

## Declarations

### Availability

All data is publicly available and summarised at https://doi.org/10.5281/zenodo.16992035. Application code is available at https://github.com/ManuelStapper/Baselines_Application.

### Contributions

S.F. conceived the study, S.F. and M.S. developed the methodology and study design, M.S. implemented the software, conducted the analysis and wrote the initial draft. S.F. and M.S. revised the manuscript. S.F. provided supervision. Both authors reviewed and approved the final version.

## Supporting Information A Data availability

**Table S1:** Availability of forecasts for COVID-19.

| Model | Availability |  | Model | Availability |  | Model | Availability |  |
| --- | --- | --- | --- | --- | --- | --- | --- | --- |
|  | States<br>(52) | Dates<br>(152) |  | States<br>(52) | Dates<br>(152) |  | States<br>(52) | Dates<br>(152) |
| AMM | 51 | 20 | IEM-MED | 52 | 68 | MUNI-ARIMA | 52 | 89 |
| BPag | 52 | 123 | IHME-Curve | 52 | 22 | MUNI-VAR | 52 | 8 |
| CEID | 52 | 93 | IQVIA | 50 | 2 | DeepSTIA | 52 | 98 |
| CH-4wk | 52 | 135 | IUPUI | 51 | 10 | OQN | 52 | 14 |
| CH-base | 52 | 152 | ISLW-STEM | 50 | 42 | PR-UMD | 51 | 21 |
| CH-ens | 52 | 135 | JBUD-HMXK | 1 | 10 | QJHong | 1 | 86 |
| CH-tens | 52 | 96 | JCB-PRM | 52 | 9 | RW-ESG | 52 | 117 |
| CH-CDCens | 52 | 75 | JHU-Bucky | 52 | 105 | SDSC-ISG | 52 | 52 |
| CU-noch | 52 | 132 | JHU-ICATTML | 52 | 3 | SigSci | 52 | 14 |
| CU-scenH | 52 | 51 | JHU-CSSE | 52 | 72 | UCF-AEM | 1 | 97 |
| CU-scenL | 52 | 130 | JHU-IDD | 52 | 114 | UCLA | 52 | 67 |
| CU-scenM | 52 | 128 | KITens | 52 | 83 | UC-C | 52 | 49 |
| CU-sel | 52 | 132 | Karlen | 52 | 106 | UMich | 51 | 70 |
| Col-SC | 1 | 55 | LANL | 52 | 65 | USACE | 52 | 18 |
| C19Sim | 52 | 56 | LNQ-ens1 | 52 | 48 | USC | 52 | 117 |
| CA-DELPHI | 52 | 103 | LU | 52 | 31 | USC-RF | 52 | 2 |
| DDS-NBDS | 52 | 35 | MIT-Cass | 49 | 82 | UVA-Ens | 52 | 96 |
| FD-Sweight | 1 | 37 | MIT-ISO | 51 | 107 | SU-GRU | 1 | 20 |
| FRBSF | 1 | 51 | MOBS | 51 | 42 | prolix | 1 | 18 |

**Table S2:** Availability of forecasts for influenza.

| Model | States<br>(52) | Availability |  |  |  | Model | States<br>(52) | Availability |  |  |  |
| --- | --- | --- | --- | --- | --- | --- | --- | --- | --- | --- | --- |
|  |  | 21/22<br>(34) | 22/23<br>(32) | 23/24<br>(31) | 24/25<br>(6) |  |  | 21/22<br>(34) | 22/23<br>(32) | 23/24<br>(31) | 24/25<br>(6) |
| CADPH | 1 | 0 | 31 | 29 | 6 | META | 1 | 0 | 0 | 0 | 5 |
| CEID | 52 | 21 | 10 | 0 | 0 | MI-J | 52 | 0 | 0 | 0 | 5 |
| CEPH | 52 | 0 | 28 | 30 | 6 | MI-E | 52 | 0 | 30 | 30 | 6 |
| CFA-F | 52 | 0 | 0 | 28 | 0 | MOBS | 52 | 20 | 31 | 30 | 5 |
| CFA-R | 52 | 0 | 0 | 30 | 0 | NEU-A | 52 | 0 | 0 | 0 | 5 |
| CMU | 52 | 24 | 31 | 28 | 6 | NEU-F | 51 | 0 | 0 | 0 | 4 |
| CU | 52 | 24 | 26 | 30 | 6 | NIH | 51 | 0 | 28 | 26 | 6 |
| FH | 52 | 0 | 0 | 30 | 4 | NU | 52 | 0 | 0 | 23 | 0 |
| FS-BL | 52 | 24 | 32 | 30 | 6 | OHT | 52 | 0 | 0 | 0 | 6 |
| FS-ENS | 52 | 24 | 31 | 30 | 6 | PSI-D | 52 | 24 | 31 | 0 | 0 |
| FS-LOP | 52 | 0 | 0 | 30 | 6 | PSI-P | 52 | 0 | 0 | 30 | 6 |
| GT-PT | 51 | 0 | 0 | 0 | 6 | PSI-PB | 52 | 0 | 0 | 20 | 4 |
| GT-PB | 51 | 0 | 0 | 0 | 6 | SG-R | 52 | 20 | 30 | 29 | 0 |
| GH-FS | 52 | 21 | 0 | 0 | 0 | SG-S | 52 | 24 | 0 | 0 | 0 |
| GH-MOD | 52 | 0 | 0 | 9 | 0 | SIG-C | 51 | 24 | 31 | 30 | 0 |
| GT-FNP | 51 | 24 | 29 | 27 | 0 | SIG-T | 51 | 24 | 31 | 30 | 6 |
| IEM | 52 | 24 | 0 | 0 | 0 | STV-GBR | 52 | 0 | 0 | 29 | 0 |
| ISUN-E | 52 | 0 | 0 | 18 | 0 | STV-ILI | 52 | 0 | 0 | 0 | 6 |
| ISUN-F | 52 | 0 | 20 | 0 | 0 | UGA-CEID | 52 | 0 | 0 | 0 | 4 |
| ISUN-G | 52 | 0 | 0 | 0 | 5 | UGA-CC | 52 | 0 | 0 | 26 | 3 |
| ISUN-N | 52 | 0 | 0 | 25 | 0 | UGA-I | 52 | 0 | 0 | 27 | 6 |
| ISUN-S | 52 | 0 | 0 | 18 | 0 | UGA-O | 52 | 0 | 30 | 3 | 0 |
| JHU-D | 52 | 0 | 0 | 0 | 6 | UGA-S | 52 | 0 | 0 | 0 | 4 |
| JHU-G | 52 | 6 | 0 | 0 | 0 | UG-CC | 52 | 0 | 0 | 28 | 4 |
| JHU-C | 52 | 0 | 0 | 17 | 4 | UG-FP | 1 | 0 | 13 | 0 | 0 |
| JHU-I | 52 | 0 | 26 | 0 | 0 | UGENS | 1 | 0 | 0 | 27 | 1 |
| LA | 52 | 24 | 23 | 28 | 3 | UI | 52 | 0 | 0 | 0 | 4 |
| LU-C | 52 | 0 | 0 | 30 | 6 | UM | 52 | 0 | 0 | 30 | 6 |
| LU-ER | 52 | 0 | 9 | 0 | 0 | UM-AR2 | 52 | 0 | 0 | 0 | 6 |
| LU-EH | 6 | 0 | 6 | 0 | 0 | UM-ARIMA | 1 | 9 | 0 | 0 | 0 |
| LU-M | 52 | 0 | 14 | 0 | 0 | UM-F | 52 | 0 | 0 | 30 | 6 |
| LU-HJ | 52 | 15 | 17 | 0 | 0 | UM-G | 52 | 0 | 9 | 0 | 0 |
| LU-HW | 52 | 0 | 12 | 0 | 0 | UM-T | 52 | 22 | 31 | 31 | 0 |
| LU-KF | 52 | 0 | 11 | 0 | 0 | UNC | 52 | 0 | 25 | 24 | 4 |
| LU-ST | 52 | 0 | 8 | 0 | 0 | UT | 52 | 24 | 0 | 0 | 0 |
| LU-TEVA | 52 | 22 | 0 | 0 | 0 | UVA-E | 52 | 24 | 30 | 29 | 6 |
| LU-V2 | 52 | 23 | 0 | 0 | 0 | VTS-ENS | 52 | 0 | 0 | 26 | 0 |
| LU-V2K | 52 | 22 | 0 | 0 | 0 | VTS-EXOG | 52 | 18 | 25 | 0 | 0 |
| LU-V2K-C | 52 | 22 | 0 | 0 | 0 | VTS-P | 52 | 0 | 0 | 0 | 6 |
| LU-V2-C | 52 | 23 | 0 | 0 | 0 | VTS-T | 52 | 0 | 5 | 0 | 0 |
| MDPRED | 1 | 0 | 0 | 0 | 6 |  |  |  |  |  |  |

## Supporting Information B PIT Functions

Figure S1 contains PIT functions of all baseline models and their corresponding CvM divergences.

**Fig. S1:**
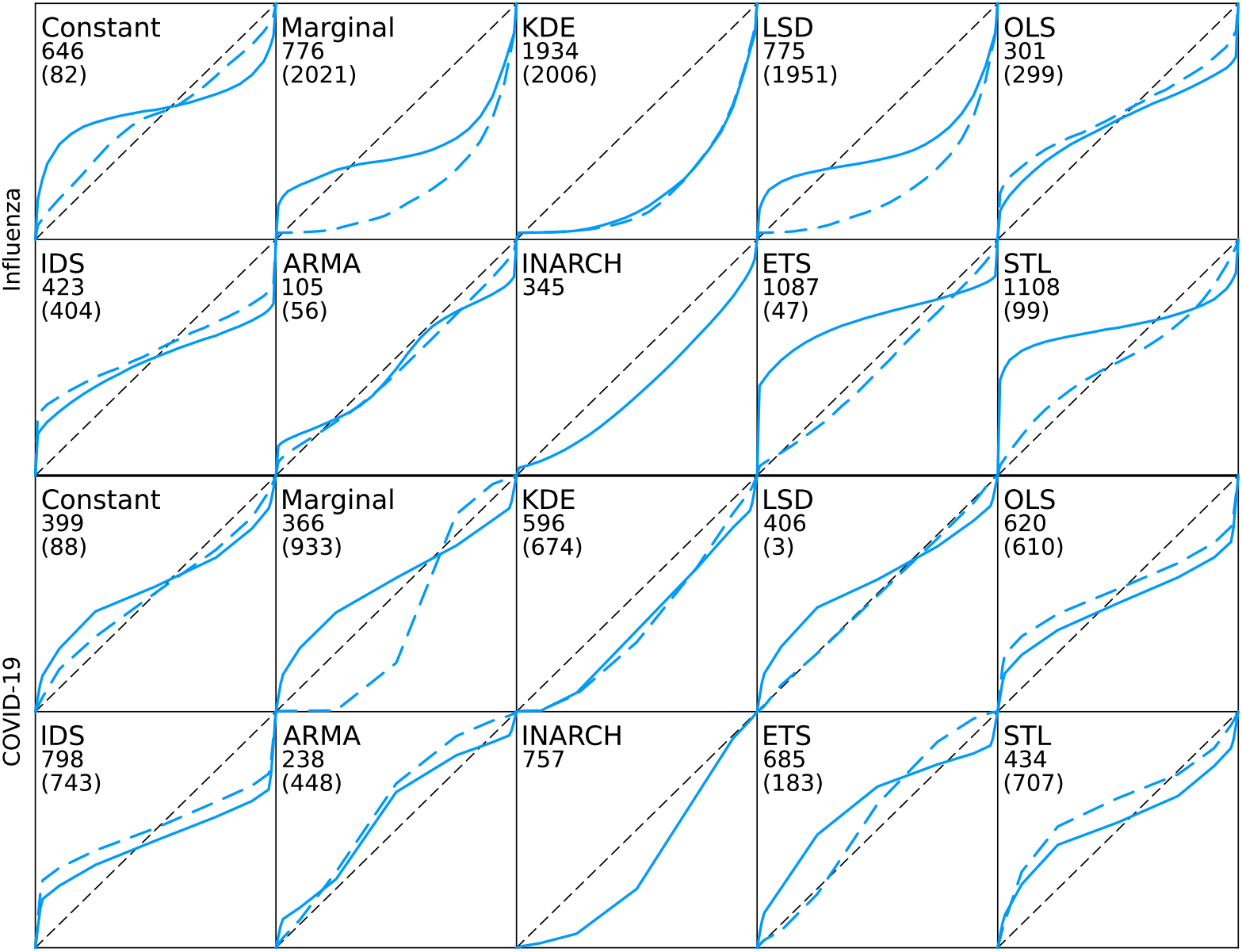
PIT functions for all baselines on original time series (solid) and log-transformed time series (dashed) - CvM divergence for baselines fitted to original time series and log-transformed time series in parentheses

